# Trust, threats, and consequences of the COVID-19 pandemic in Norway and Sweden – a comparative survey

**DOI:** 10.1101/2020.05.16.20089953

**Authors:** Lise M. Helsingen, Erle Refsum, Dagrun Kyte Gjøstein, Magnus Løberg, Michael Bretthauer, Mette Kalager, Louise Emilsson, for the Clinical Effectiveness Research group

**Author notes:** Lise M. Helsingen and Erle Refsum contributed equally to this article. Mette Kalager and Louise Emilsson contributed equally to this article. **Corresponding author:** Lise M. Helsingen, Clinical Effectiveness Research, Department of Health Management and Health Economics, University of Oslo, PO Box 1089 Blindern, 0318 Oslo.

## Abstract

**Objectives:** Norway and Sweden have similar populations and health care systems, but different reactions to the COVID-19 pandemic. Norway closed educational institutions, and banned sports and cultural activities; Sweden kept most institutions and training facilities open. We aimed to compare peoples’ attitudes towards authorities and control measures, and effects on life in Norway and Sweden.

**Design:** Anonymous web-based surveys for individuals age 15 or older distributed through Facebook using the snowball method.

**Setting:** Norway and Sweden, mid-March to mid-April, 2020.

**Participants:** 3,508 individuals participated in the survey (Norway 3000; Sweden 508). 79% were women, the majority were 30-49 years (Norway 60%; Sweden 47%), and about 45% of the participants in both countries had more than four years of higher education.

**Outcome measures:** Perceived threat of the pandemic, views on infection control measures, and impact on daily life. We performed descriptive analyses of the responses and compared the two countries.

**Results:** Participants had high trust in the health services, but differed in the degree of trust in their government (High trust in Norway 17%; Sweden 37%). More Norwegians than Swedes agreed that school closure was a good measure (Norway 66%; Sweden 18%), that countries with open schools were irresponsible (Norway 65%; Sweden 23%), and that the threat from repercussions of the mitigation measures were large or very large (Norway 71%; Sweden 56%). Both countries had a high compliance with infection preventive measures (> 98%). Many lived a more sedentary life (Norway 69%; Sweden 50%) and ate more (Norway 44%; Sweden 33%) during the pandemic.

**Conclusion:** Sweden had more trust in the authorities, while Norwegians reported a more negative lifestyle during the pandemic. The level of trust in the health care system and self-reported compliance with preventive measures was high in both countries despite the differences in infection control measures.

**Strengths and limitations of this study:**

- This study compares people’s attitudes to the handling of the COVID-19 pandemic in Norway and Sweden, two very similar countries with different strategies
- The study provides a situation assessment of the attitudes and self-reported compliance to infection preventive measures taken by Norwegian and Swedish authorities
- The study participants were recruited to answer the survey through Facebook and their views may not be generalizable to the whole population in Norway and Sweden
- The surveys were distributed three weeks apart in time in the two countries. The situation was stable during that time-period in both countries.

## Introduction

Since the first detected case of COVID-19 in Norway and Sweden in mid-February 2020, the public health authorities in the two neighboring countries have provided advice on well-known infection control measures, such as hand hygiene, sneezing and coughing habits, isolation of individuals with COVID-19 symptoms, and tracing of contacts of confirmed cases. Further, people are advised to avoid unnecessary travels and to work from home if possible. Such measures represent a relatively small burden to the societies.

On March 12, Norway issued stricter measures and instituted quarantine for those who entered the country. The same day, the government closed all kindergartens and schools, physiotherapists, psychologists, hairdressers, swimming pools and training centers, banned all sporting and cultural events and all organized sports, including children’s sports (1). Sweden chose a different strategy: Kindergartens, elementary schools, training facilities and other businesses were kept open, and children’s sports continued. High schools and universities were closed on March 18, and on March 29, the Swedish government applied a general rule against assemblies of more than 50 people (2). Sweden did not restrict border crossing. In both countries, people above 70 years were advised to limit social contact and stay at home if possible.

Norway and Sweden have similar ethnic and sociodemographic profiles and age distributions of the population, and similar health care, educational and political systems (3, 4). The countries have similar cultures and languages, and understand each other and can follow each other’s media and public announcements. We were curious about the perception of threat due to the COVID-19 pandemic and people’s trust in authorities’ crisis management in the two countries, and conducted population surveys in Norway and Sweden from mid-March to mid-April 2020.

## Methods

Based on an ongoing COVID-19 project with focus group interviews in the Norwegian population, we designed a survey to investigate people’s attitudes and opinions about infection control measures during the pandemic, their trust in the government and health care, and changes in daily life that may affect public health, in Norway and Sweden. We tested the survey on 10 Norwegian volunteers, and revised it based on their input. Although Norwegian and Swedish languages are closely related, and most people understand the other language well, a native speaking Swede translated the Norwegian version of the survey to Swedish to avoid any misunderstandings. We used the Norwegian version for Norwegian participants, and the Swedish version for Swedish participants. The questions were identical in the Norwegian and Swedish version, except for a few country-specific questions.

The questionnaire covered the following COVID-19 related topics: Perceived threat from the pandemic, trust in the authorities, opinions about infection control measures, solidarity and social control, and changes in daily life during the pandemic. We also asked participants about sex, age, educational level and municipality of residence.

Most questions consisted of statements in which respondents rated how much they agreed or disagreed on a 6-point Likert scale, stating 1 “Strongly disagree” and 6 “Strongly agree” (5). We did not include a neutral category as we observed that people did not have neutral opinions in the focus group interviews. Instead, we applied a “do not know” category in statements about threat. Appendix 1 displays the entire surveys in Norwegian and Swedish, with all questions and responses in both countries.

We used the University of Oslo’s system for digital data collection (6). The study was anonymous and did not require approval by the Norwegian Center for Research Data or ethics committee for medical research, or the Swedish Ethical Review Authority (7, 8). The focus group interview study that informed the questionnaire was approved by the Norwegian Center for Research Data.

We disseminated a link to the questionnaire through the investigators’ Facebook pages using the “snowball method”, to persons 15 years or older in each country (9). We asked individuals to share the survey (request) with others in their networks.

We decided to close the survey when reaching at least 500 participants in each country. With 500 participants, the margin of error is about 4.5% around the point estimates, which we considered to be sufficiently accurate for our purpose (10).

We received responses in Norway from March 20-21 and in Sweden from April 10-15. Participants had to respond to all questions to be able to submit the survey. There was no time limit for completing the survey.

We analyzed the data descriptively, using Stata version 16.1 (StataCorp, College Station, Texas, United States). For analyses, we compared the distribution of responses on the 6-point Likert rating scale in the two countries. We did not have predefined hypotheses about potential differences between Norway and Sweden or between subgroups of the study populations, and did not perform statistical significance tests. We analyzed all responses to check if there was variation related to sex, age, municipality size, or educational level, and report where large differences between these subgroups were observed.

### Patient and public involvement

Ten Norwegian volunteers, recruited through the authors’ networks, tested the first version of the Norwegian survey and commented regarding the understanding of the questions, time required to participate in the survey, and if they missed important topics. The survey was revised based on their comments before it was distributed to the public.

## Results

We received 3,508 responses: 3,000 from Norway and 508 from Sweden. Demographics of participants were similar in the two countries (table 1).

**Table 1:** Demographics

|  |  | Norway | Sweden |
| --- | --- | --- | --- |
|  |  | N (%) | N (%) |
| Responders |  | 3,000 | 508 |
| Age |  |  |  |
|  | 15-29 | 390 (13) | 58 (11) |
|  | 30-49 | 1,768 (59) | 238 (47) |
|  | 50-64 | 710 (24) | 159 (31) |
|  | 65 and older | 132 (4) | 53 (10) |
| Sex |  |  |  |
|  | Women | 2,350 (78) | 412 (81) |
|  | Men | 632 (21) | 91 (18) |
|  | Not reported | 18 (<1) | 5 (1) |
| Educational level* |  |  |  |
|  | Elementary school or upper secondary school | 81 (3) | 4 (1) |
|  | High school | 723 (24) | 125 (23) |
|  | Higher education <4 years | 830 (28) | 148 (29) |
|  | Higher education >4 years | 1,366 (46) | 231 (45) |
|  | Currently attending school or university | 354 (12) | 51 (10) |
| Number of inhabitants in the residing county |  |  |  |
|  | Rural areas<sup>1</sup> | 1,606 (54) | 391 (77) |
|  | Urban areas<sup>2</sup> | 1,264 (42) | 109 (22) |
|  | Unkown county | 130 (4) | 8 (2) |
| Population |  | 5 300 000 | 10 100 000 |

\* Highest completed education
<sup>1</sup> Defined as number of inhabitants less than or equal to 100, 000 in Norway and 200, 000 in Sweden
<sup>2</sup> Defined as number of inhabitants 100,000 and more in Norway and 200,000 and more in Sweden

### COVID-19 infection preventive measures and trust

The Swedish participants placed a higher level of trust in their government and health authorities than the participants in Norway (In Norway 17% and 24% strongly agreed they trusted their government and health authorities, respectively, and in Sweden 37% and 50%, respectively) (figure 1). Most people trusted the health services and hospitals to similar degree in both countries (figure 1).

**Figure 1:**
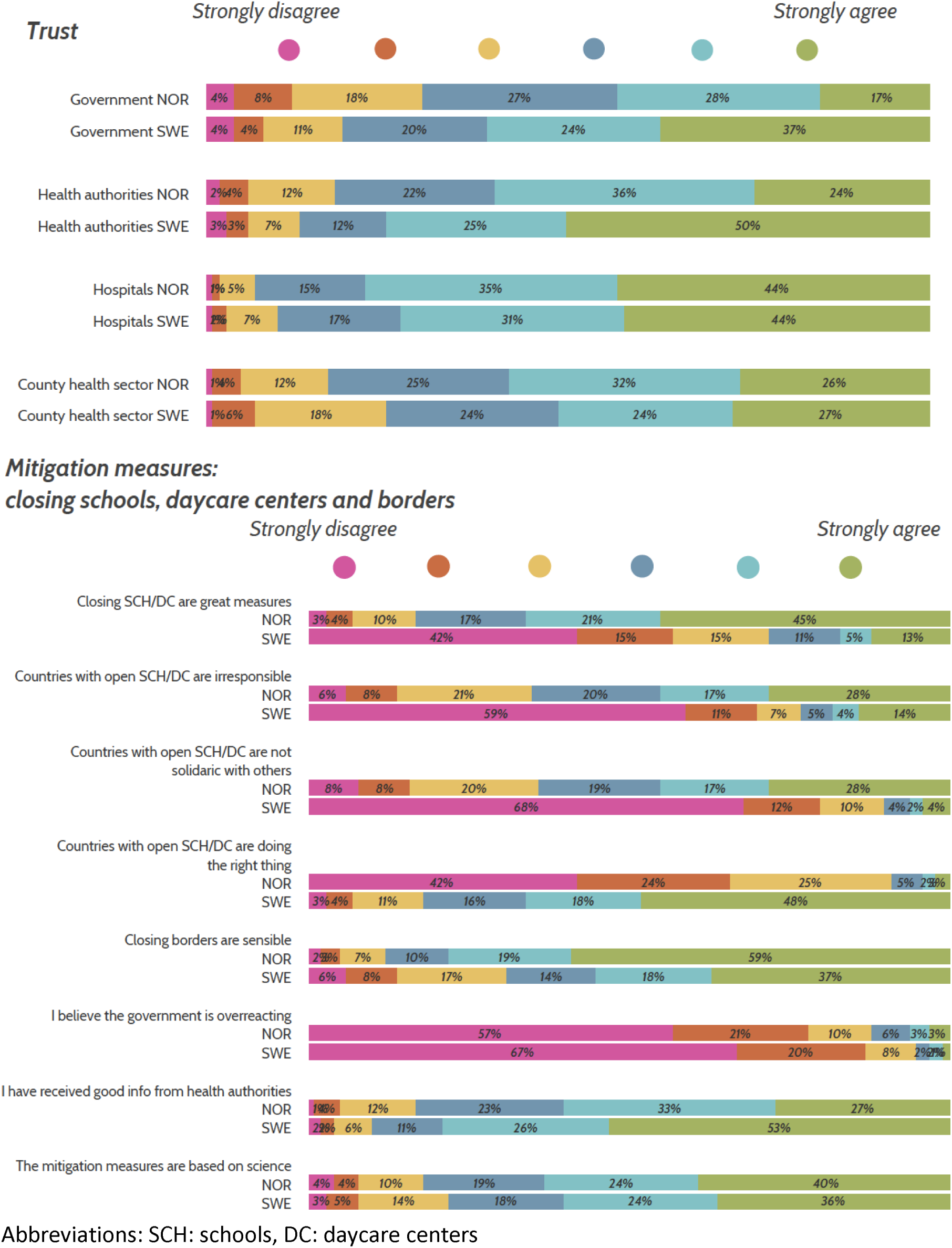
Trust in institutions and mitigation measures in Norway (NOR) and Sweden (SWE). The proportion (%) of responders who disagreed or agreed.

In Sweden, 53% strongly agreed that they received good information from their health authorities during the pandemic, compared to 27% in Norway (figure 1).

Most Swedes disagreed that closing schools and kindergartens were great measures (42% strongly disagreed), that countries with open schools and kindergartens are irresponsible (59% strongly disagreed), and that keeping these institutions open represented lack of solidarity with other countries (68% strongly disagreed). Most Norwegians, on the other hand, agreed that closing of schools and kindergartens were great measures (45% strongly agreed), that countries with open schools and kindergartens are irresponsible (28% strongly agreed), and that it was a lack of solidarity with other countries to keep them open (28% strongly agreed). Only 3% of Norwegians strongly agreed that countries keeping school and kindergartens open were doing the right thing, whereas 48% of the Swedes strongly agreed. The majority in both countries strongly disagreed that their government was overreacting (57% in Norway and 67% in Sweden) (figure 1).

A majority in both countries agreed that the authorities’ decisions on infection preventive measures were based on scientific evidence (83% in Norway and 78% in Sweden agreed) (figure 1).

The compliance with infection preventive measures was high and similar in the two countries, but more people worked from home to protect themselves and others in Norway than in Sweden (table 2).

**Table 2:** Number and amount of individuals that responded that they took the following measures to protect themselves and others from COVID-19 in Norway and Sweden

|  | Protect yourself |  | Protect others |  |
| --- | --- | --- | --- | --- |
|  | Norway (%) | Sweden (%) | Norway (%) | Sweden (%) |
| Work from home | 1,759 (59) | 213 (42) | 1,751 (58) | 213 (42) |
| Avoid people that cough | 2,126 (71) | 362 (71) | * | * |
| Cough in elbow hook or disposable handkerchief | * | * | 2,564 (85) | 438 (86) |
| Hand-wash | 2,957 (99) | 486 (96) | 2,947 (98) | 498 (98) |
| Use gloves | * | * | 832 (28) | 103 (20) |
| Use face mask | 86 (3) | 7 (1) | 105 (4) | 13 (3) |
| Do not meet friends | 2,600 (87) | 279 (55) | 2,619 (87) | 297 (58) |
| Avoid public transportation | 2,472 (82) | 366 (72) | 2,494 (83) | 381 (75) |
| Stay home during my spare time | 2,622 (87) | 362 (71) | 2,644 (88) | 377 (74) |
| No special measure | 11 (0.4) | 8 (2) | 9 (0.3) | 3 (1) |

\* Did not ask

### COVID-19 as a health threat

In Norway, 53% responded that COVID-19 is a large to very large threat to the population, compared to 58% in Sweden (figure 2). More people in Norway (41%) than in Sweden (21%) believed that the virus posed a large to very large health threat to their family members. A minority (2-3%) in both countries, thought that the virus poses a very large health threat to them personally, but this varied largely with age: While only 10% and 9% of young adults between the ages 15-29 in Norway and Sweden, found that the virus posed a large or very large threat to them, 38% and 15% of people 65 years or older believed the coronavirus posed a large or very large threat in Norway and Sweden, respectively.

More than 60% of both Norwegians and Swedes agreed that the economic crisis would be a larger challenge than the pandemic itself. About half of the participants in both countries worried about their personal economy and 71% of the Norwegians thought that the ripple effects of the preventive measures against the coronavirus represent a large or very large threat to their society, compared to 56% of the Swedes (figure 2).

**Figure 2.**
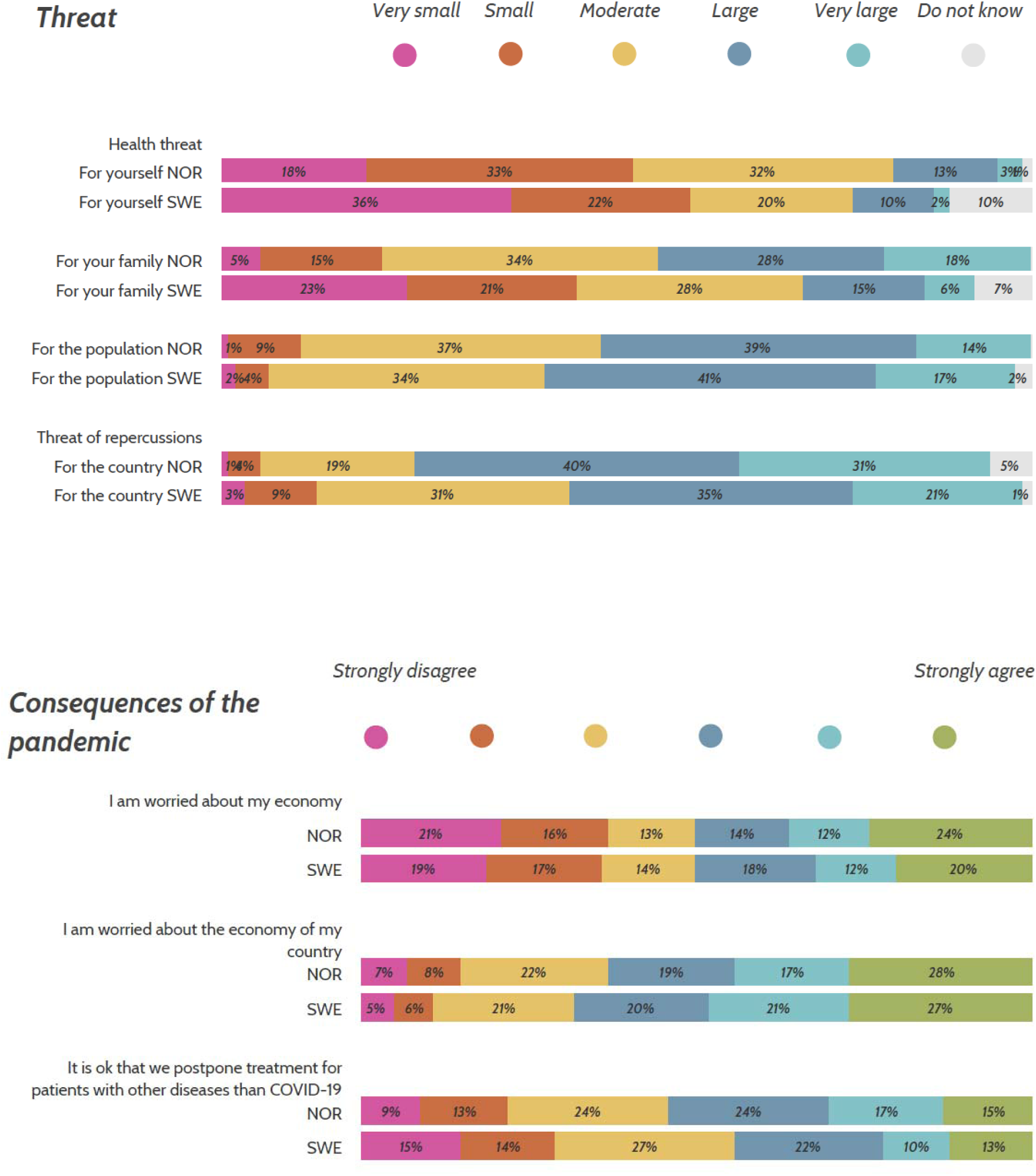
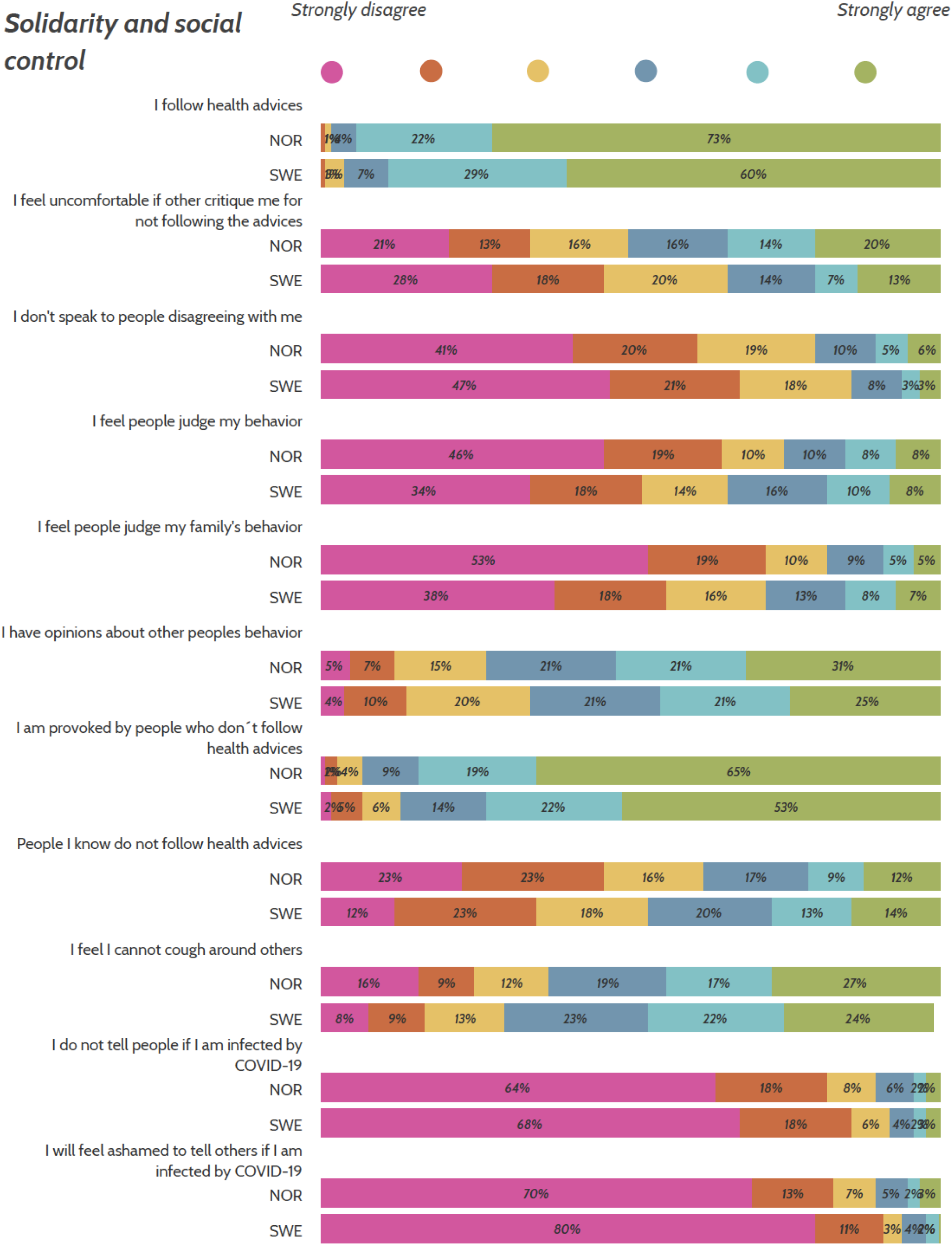
COVID-19 as a threat, the consequences of the pandemic and solidarity and social control in Norway (NOR) and Sweden (SWE). The proportion (%) of responders who thought COVID-19 was a very small to very large threat, and the proportion of responders who strongly disagreed or agreed to the consequences, solidarity and social control.

### Daily life and lifestyle

People reported high compliance with general infection prevention measures such as hand hygiene (>95% in both countries) and cough habits (>85% in both countries) (table 2). Most Norwegians (88%) and Swedes (74%) stayed at home during their spare time, but a larger proportion of Norwegians (87%) than Swedes (58%) reported that they do not meet friends (table 2). In both countries, the majority of people were provoked by people who did not follow the advice from the authorities (Norway 84%, Sweden 75%), whereas 16% of Norwegians and 34% of Swedes felt that others were judging their behavior related to the pandemic (figure 2).

In Norway, 69% lived a more sedentary life during the pandemic versus 50% in Sweden, and 44% in Norway ate more versus 33% in Sweden (figure 3). More people were depressed and sad in Norway than in Sweden (21% and 41% in Norway, 15% and 18% in Sweden, respectively). Around 60 % in both countries felt that their life were put on hold, and at the same time felt useful, and more than 70% felt proud of how they coped with the situation (figure 3).

There were no major differences in the results when stratifying by age groups, sex, municipality size and educational level. Appendix 1 displays the entire survey in Norwegian and Swedish, and all responses.

**Figure 3.**
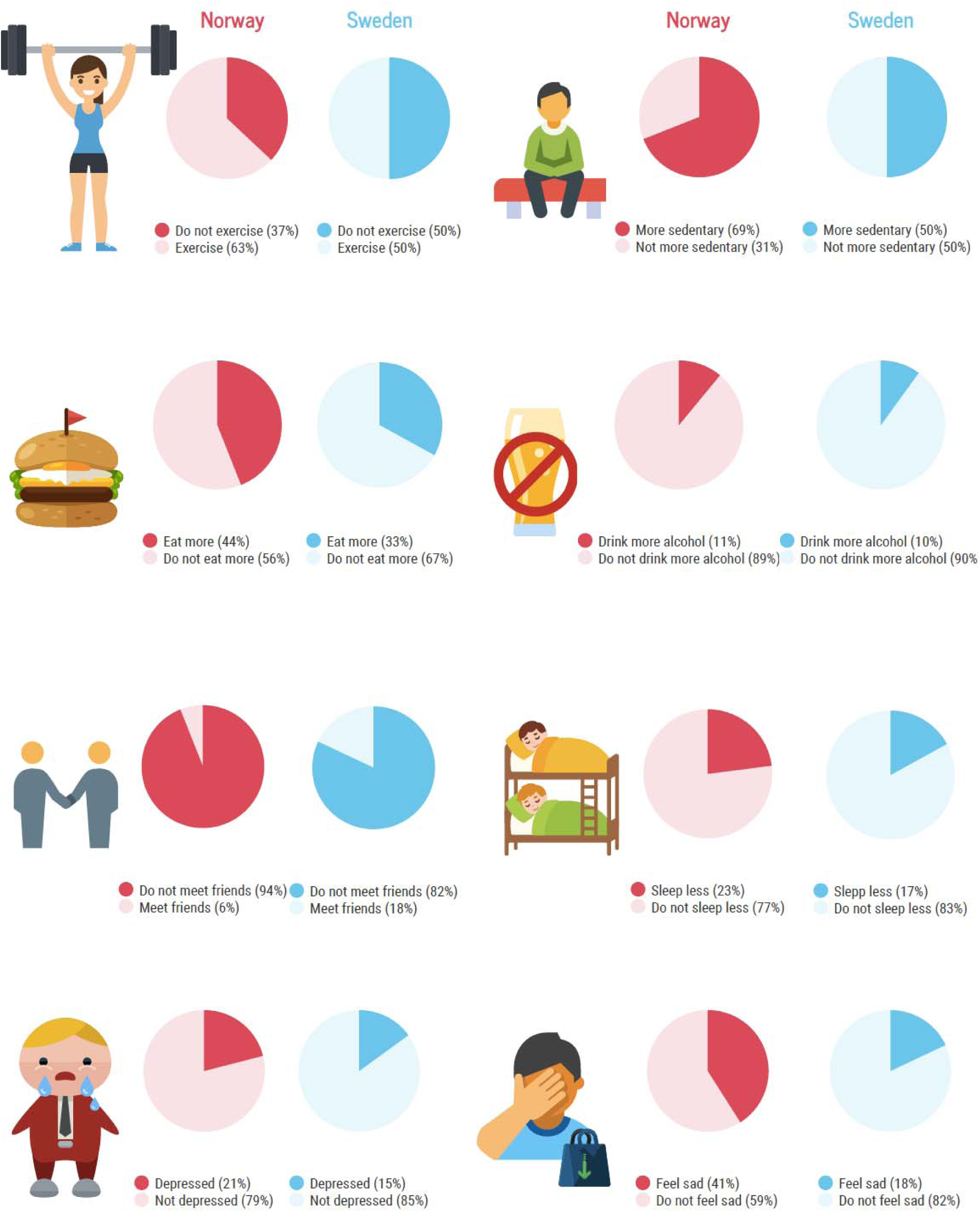
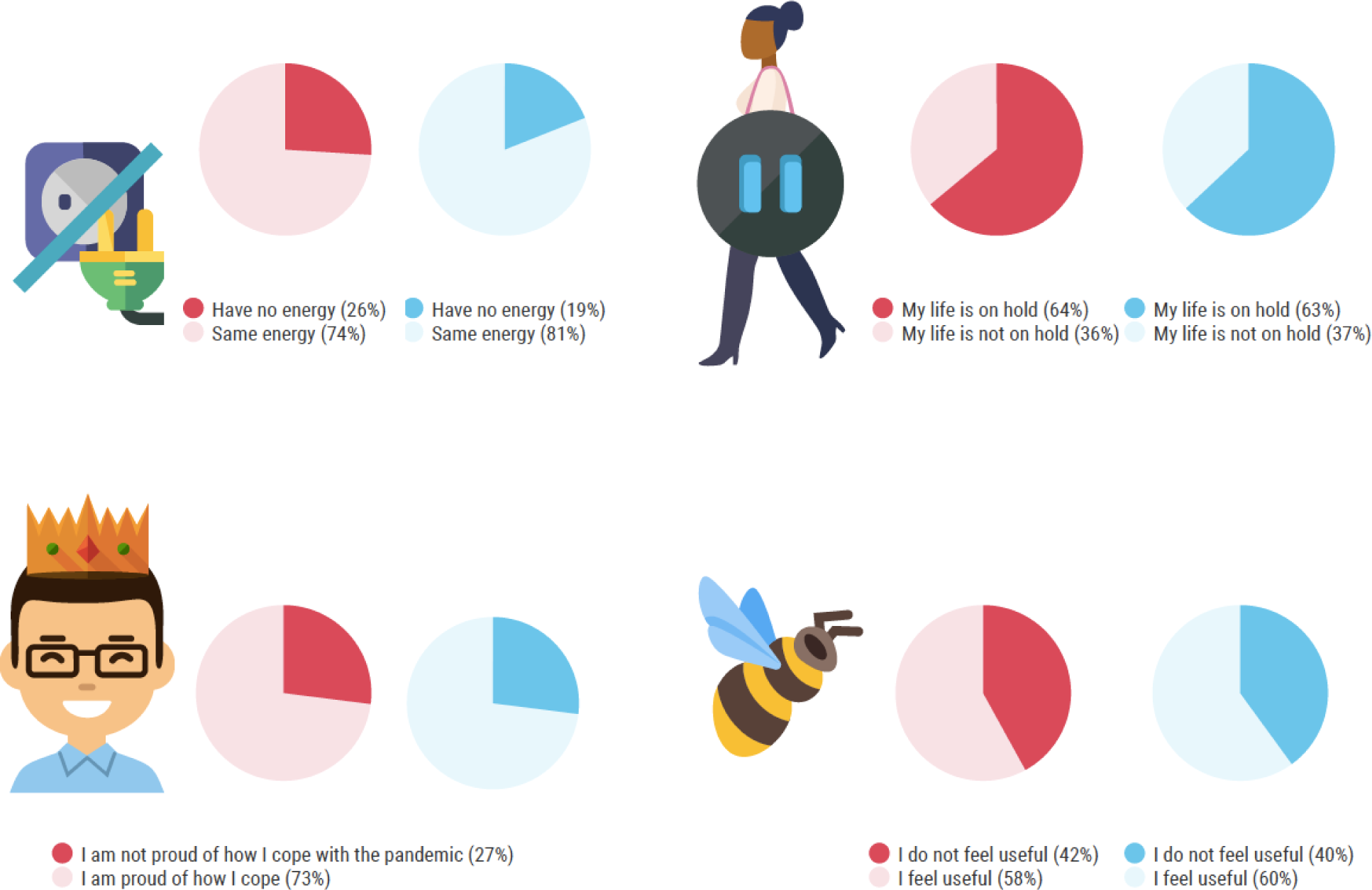
Changes in daily habits during the first phases of the COVID-19 pandemic in Norway and Sweden. The dark color in the pie-chart indicate habit that may influence public health negatively.

### Discussion

This survey provides a snapshot of people’s opinions, fears and attitudes towards preventive measures during the COVID-19 pandemic in Norway and Sweden in March and April 2020. People had a high degree of trust in the health systems and authorities in both countries. Although the countries’ authorities have taken different approaches to the pandemic, most people support their authorities’ COVID-19 preventive measures. The large majority of people respond that they comply with the advice or regulations given by their governments and health authorities, and are provoked by people who do not comply.

### Trusty neighbors

Norway and Sweden are neighboring countries, and have a long history of close collaboration. Many Norwegians study and work in Sweden and vice versa. The culture and the languages are very similar. Both countries are parliamentary democracies with a strong social democratic profile: Both have mainly public health care systems, solid social welfare programs, and all education, including university, is free of charge. Despite the similarities during normal circumstances, the two countries have responded differently to the COVID-19 pandemic. The main difference is the closing of borders and the closure of schools and kindergartens and different businesses in Norway, which remained open in Sweden.

Our results indicate that there is a high degree of trust in government and health services in both Norway and Sweden. Few disagree with their governments’ measures or were provoked by them, even though the preventive measures varied substantially between two countries. Most of the Norwegian participants felt that the closure of kindergartens and schools were appropriate, while in Sweden, most people thought it was inappropriate to close schools and kindergartens.

The Nordic countries have the highest levels of generalized trust and social capital in the Western World under normal circumstances, and these high levels have been stable for decades (11, 12). Social capital can be defined as “the ability to cooperate without written rules and extensive contracts” (13), and may therefore be linked to a population’s commitment to follow guidance from the authorities. Both Norway and Sweden have a long history of successful volunteer infection preventive measures, such as child vaccinations, where both countries reach a coverage of more than 95% (14, 15). According to both countries laws – volunteer preventive measures should be applied first, whenever possible (16, 17).

According to our survey, the Norwegians and the Swedes seems so far during the pandemic to be loyal to their authorities, and trust that their respective governments’ decisions are based on scientific evidence. In both countries, people reported a high level of compliance to social distancing and hygiene, although these measures have been introduced through regulations in Norway in contrast to guidance in Sweden.

More than 70% of the study population in both countries are proud of how they cope with the situation. Thus, our results indicate that both the Norwegian and Swedish government’s call to people’s solidarity and to join forces as a national virtue, seem to work so far.

### Health Effects

Over the last months, the people in Norway and Sweden have been told to work from home if possible, and to limit all travel. In Norway, people travelling into the country are mandated a 14 day quarantine (18). Many people are laid off work, and as of April 21, 2020, the level of unemployment is 10.2% in Norway, and 15% are seeking employment (19), the highest number since Second World War. In Sweden, the proportion of unemployed has to date increased only slightly compared to previous years (20, 21). In Norway, all sports and cultural events are banned and gyms are closed, while in Sweden, gyms and training facilities are open and organized children’s sports arrangements are encouraged, based on a judgment that the benefit of socializing and being physically active outweighs the potential risks of COVID-19 for children. The Swedish Communicable Diseases Act specifies that when infection preventive measures affects children, particular attention must be paid to what the child’s best interests require (16), whereas no similar statement is found in the Norwegian law (17).

It is natural to think that the consequences of the pandemic and the preventive measures can have a negative impact on people’s lives and health (22). In our survey, more than 80% in both countries say that they do not live their lives as usual. Some ate more, many are exercising less, and some drink more alcohol than they do usually. Many feel that their lives are on hold and many people, especially in Norway, feel sad and depressed. Possible negative health consequences as a result of measures to prevent the spread of the coronavirus must be taken into account when considering the duration and usefulness of infection prevention measures.

### Strengths and limitations

Our study sample was collected using the snowball method and the responses may not be generalizable to the general population in Sweden and Norway. The method is frequently used in surveys, especially when there is a need to quickly document a situation that changes over time (9). Our sample did not include children below 15 years of age, and we had few participants below 29 and 70 years and older. The majority of the participants were women, and we had an overrepresentation of the age group 30 to 49 and of people with higher education, compared to the general population (23).

The survey was first performed in Norway, about ten days after the Norwegian government had introduced the most restrictive infection prevention measures ever seen in Norway. The media focus and the population’s attention to the pandemic was large at the time. This resulted in 3000 responses in less than 24 hours. In Sweden, the survey was performed three weeks later, and we needed six days to reach our goal of 500 respondents. This may have influenced the comparability between the two countries. However, during the entire study period from mid-March to mid-April 2020, the COVID-19 situation was stable in both countries, and no substantial new preventive measures were taken in this period, although the COVID-19 mortality rates increased more in Sweden than in Norway (24). We repeated the survey in Norway in the period April 4th-8th and found that the results did not differ substantially from the results from the first round presented here (appendix 2). In addition, other surveys performed repetitively in the population shows that the level of trust in the government was stable and high in Norway in the period between March 15 and 31 (25). A survey performed repetitively in Sweden also shows that the trust in government and health care system was high and stable from March 21 to April 8 (26). Thus, we believe that our survey provides a valid comparison of the two countries.

### Conclusion

Our results show that both people in Norway and Sweden have a high level of trust in their government, despite different handling of the COVID-19 pandemic. The authorities in both countries experience a high level of compliance and acceptance to infection prevention measures from their populations. This indicates that preventive measures can be applied successfully both through regulations, like in Norway, and through advice based on mutual trust between the authorities and the population, like in Sweden. Basing infection prevention on volunteer measures has been a long-standing tradition and is a statutory right in both countries, and has probably been the recipe for success for instance to achieve a high vaccine coverage (14, 15). Danish scientists wrote in 2015 about the stock of social trust being an important part of the reason for the successful Scandinavian welfare-state (27). They end their piece with a warning: “If the Scandinavian high-trust societies should in the future turn into control societies, they will probably no longer be among the world’s leading countries in terms of socio-economic success” (12).

## Data Availability

Data will be shared to interested parties. Please contact professor Mette Kalager

## Author contributions

LMH, MK, DKG and ML contributed to the conception and design of the work, and all authors contributed to the acquisition and interpretation of data; ER, LE and LMH contributed to the analysis of the data; LMH and MB drafted the paper and all authors revised it critically, approved the final version and are accountable for all aspects of the work. LMH and ER, and MK and LE contributed equally to this paper.

## Data sharing

Data will be shared to interested parties. Please contact Prof. Mette Kalager.

## Ethics approval

This was an anonymous survey, with no link between researchers and participants, and the answers can in no way be linked to identifying information about the respondent. According to a ruling by the Regional Committee for Medical and Health Research Ethics (REC), this research is outside the remit of REC and does not need an approval from REC. (https://helseforskning.etikkom.no/reglerogrutiner/soknadsplikt/sokerikkerek?p_dim=34999).

## Transparency statement

LMH affirms that this manuscript is an honest, accurate, and transparent account of the study being reported; that no important aspects of the study have been omitted; and that any discrepancies from the study as planned have been explained.

## Dissemination to participants and related patient and public communities

We will disseminate the results on Clinical Effectiveness Research Group’s web page: https://www.med.uio.no/helsam/forskning/grupper/klinisk-effektforskning/, as well as in newspapers and social media.

## Exclusive licence

I, the Submitting Author has the right to grant and does grant on behalf of all authors of the Work (as defined in the below author licence), an exclusive licence and/or a non-exclusive licence for contributions from authors who are: i) UK Crown employees; ii) where BMJ has agreed a CC-BY licence shall apply, and/or iii) in accordance with the terms applicable for US Federal Government officers or employees acting as part of their official duties; on a worldwide, perpetual, irrevocable, royalty-free basis to BMJ Publishing Group Ltd (“BMJ”) its licensees and where the relevant Journal is co-owned by BMJ to the co-owners of the Journal, to publish the Work in BMJ Open and any other BMJ products and to exploit all rights, as set out in our <u>licence</u>.

The Submitting Author accepts and understands that any supply made under these terms is made by BMJ to the Submitting Author unless you are acting as an employee on behalf of your employer or a postgraduate student of an affiliated institution which is paying any applicable article publishing charge (“APC”) for Open Access articles. Where the Submitting Author wishes to make the Work available on an Open Access basis (and intends to pay the relevant APC), the terms of reuse of such Open Access shall be governed by a Creative Commons licence – details of these licences and which <u>Creative Commons</u> licence will apply to this Work are set out in our licence referred to above.

